# Objectively measured social media use and psychosocial wellbeing among adolescent girls: a prospective study

**DOI:** 10.64898/2026.05.25.26354016

**Authors:** Silja Kosola, Sara Mörö, Elina Holopainen

## Abstract

**Objective:** Cross-sectional studies indicate associations between self-reported social media use and adolescent wellbeing outcomes. We aimed to evaluate longitudinal associations of objectively measured smartphone and social media use with psychosocial wellbeing.

**Design:** Observational study with one year of follow-up

**Setting:** High schools in Finland from 2022 to 2023

**Population:** 259 adolescent girls (mean age 16.3 years at baseline)

**Main outcome measures:** screenshots depicting smartphone and social media use, Bergen Social Media Addiction Scale (BSMAS), Generalized Anxiety Disorder-7 questionnaire, Body Appreciation Scale 2 (BAS-2) and visual analogue scales (VAS) of mood, tiredness, and loneliness

**Results:** Across one year of follow-up, anxiety, body appreciation, and mood improved, but possible social media addiction increased from 15% to 17%. Social media addiction at baseline was associated with increased anxiety (r=0.29, p<0.001), lower body appreciation (r=-0.15, p=0.022), and more loneliness (r=0.20, p=0.001) at follow-up. Anxiety at baseline was associated with social media addiction at follow-up (r=0.19, p=0.005). The highest quartile of TikTok users reported more social media addiction (BSMAS 19 [IQR 16-21] vs. 17 [IQR 14-20]; p=0.009) and lower body appreciation (BAS-2 32 [IQR 28-38] vs. 35 [IQR 29-40]; p=0.003) than did others. The highest quartile of Snapchat users reported more social media addiction (BSMAS 19 [IQR 15-21] vs. 17 [IQR 14-20]; p=0.007) and tiredness (VAS 21 [IQR 13-32] vs. 26 [IQR 15-35]; p=0.049) than did others.

**Conclusions:** Consistent with cross-sectional studies, social media addiction was associated with poorer psychosocial outcomes across follow-up. Policies to protect adolescents from social media addiction are urgently needed.

**Key messages:** *What is already known on this topic:* Longitudinal studies utilizing objectively collected data on smartphone and social media use to assess associations with adolescent wellbeing are infrequent, as are detailed analyses regarding different social media platforms.

*What this study adds:* The bidirectional association between social media addiction and anxiety across one year of follow-up suggests the possibility of a vicious cycle. The most active users of TikTok and Snapchat scored especially high on social media addiction but presented some differences in wellbeing profiles.

*How this study might affect research, practice or policy:* Screenshots appear to be a valid instrument for objective assessment of smartphone and social media use. Policies are urgently needed to limit the addictiveness of social media.

## Introduction

Smartphones and social media have become nearly ubiquitous in the daily lives of adolescents, affecting how young people communicate, entertain themselves, and construct social identities (1). Recent epidemiological and survey studies show high smartphone use and social media engagement, with average daily use measured in hours (2-4). The change has been rapid: a decade ago, most adolescents used social media for less than 30 minutes daily (5).

Concurrently with the increased use of social media, epidemiological studies have reported a rise in adolescent mental health problems, especially among girls (6,7). Concerns about the potential harms of social media to adolescent mental health and wellbeing are based on several plausible mechanisms. Social comparison and feedback-seeking are central online behaviors that can amplify fear of missing out (FOMO), anxiety, and body-image concerns (8,9). These processes are particularly salient during mid-adolescence and may disproportionately affect girls. Indeed, girls tend to show symptoms of social media addiction more frequently than boys, who in turn may develop digital gaming addiction more frequently than girls (10,11). If children and adolescents of different ages and genders are included in studies, developmental and gender differences may be diluted.

Despite the wealth of studies conducted on the associations of social media and adolescent mental health, most studies are cross-sectional, thus impeding assessment of directionality (12,13). Longitudinal studies tend to rely on self-reported smartphone and social media use instead of objectively measured data (2,14). When objective data and self-reports on screen time have been compared, the correlation has been moderate (4). Some longitudinal studies also date to the era prior to modern algorithms, which employ many psychological mechanisms associated with behavioral addictions and which have made social media platforms more compelling than they were a decade ago (15,16). This evolution of social media platforms also means that their effects on adolescent wellbeing may differ from one another. To date, only a few studies have focused on the use of specific social media platforms, most often TikTok (17).

We aimed 1) to estimate directionality between social media addiction and psychosocial wellbeing, and 2) to evaluate the differences between the most popular social media applications and associated psychosocial wellbeing using longitudinally collected objective data on social media use. We hypothesized that bidirectional associations between social media addiction and psychosocial wellbeing would emerge and that psychosocial outcomes might differ depending on the social media platforms used.

## Methods

### Setting and design

The School, Sport and Social media study (3S) is a prospective study conducted in the capital region of Finland (Helsinki, Espoo, and Vantaa; background population 1.2 million).

After developing the study documents with the nine members (ages 15-17) of the Youth Research Advisory Board of the Helsinki University Hospital (Helsinki, Finland), we obtained ethics approval from the HUS Regional Committee on Medical Research Ethics (HUS/117/2022) and research permits from the respective municipal educational administrations. Principals of 21 geographically and socioeconomically diverse high schools gave permission to inform students about the study. Please see reference 4 for details.

### Study population

In autumn 2022, 1164 adolescent girls completed study surveys and 564 of them (48 %) also sent screenshots depicting their smartphone use during the past week. The baseline results have been published previously (4). The baseline population was representative of female high school students in the study area. One year later in autumn 2023, a link to the follow-up survey was sent via email to the original study cohort. At both times, responses were collected using RedCap, a secure online tool. After each response, participants received a movie ticket (value 9€).

### Smartphone and social media use

Participants were asked to estimate their daily smartphone use and to attach screenshots of their smartphone use (iPhone Screen Time, Huawei Digital Balance, Android Digital Wellbeing) from the past week. From the screenshots, we collected the following data: number of days with data available, names of most frequently used applications, and time spent using each of them. Total screen time was divided by the number of days with data available to measure average daily smartphone use. We summed the time spent using similar types of applications (e.g. social media, streaming) and divided this sum by the number of days with data available to estimate average daily time spent on different activities. Please see supplemental table 1 of our baseline publication for details (4).

**Table 1.**
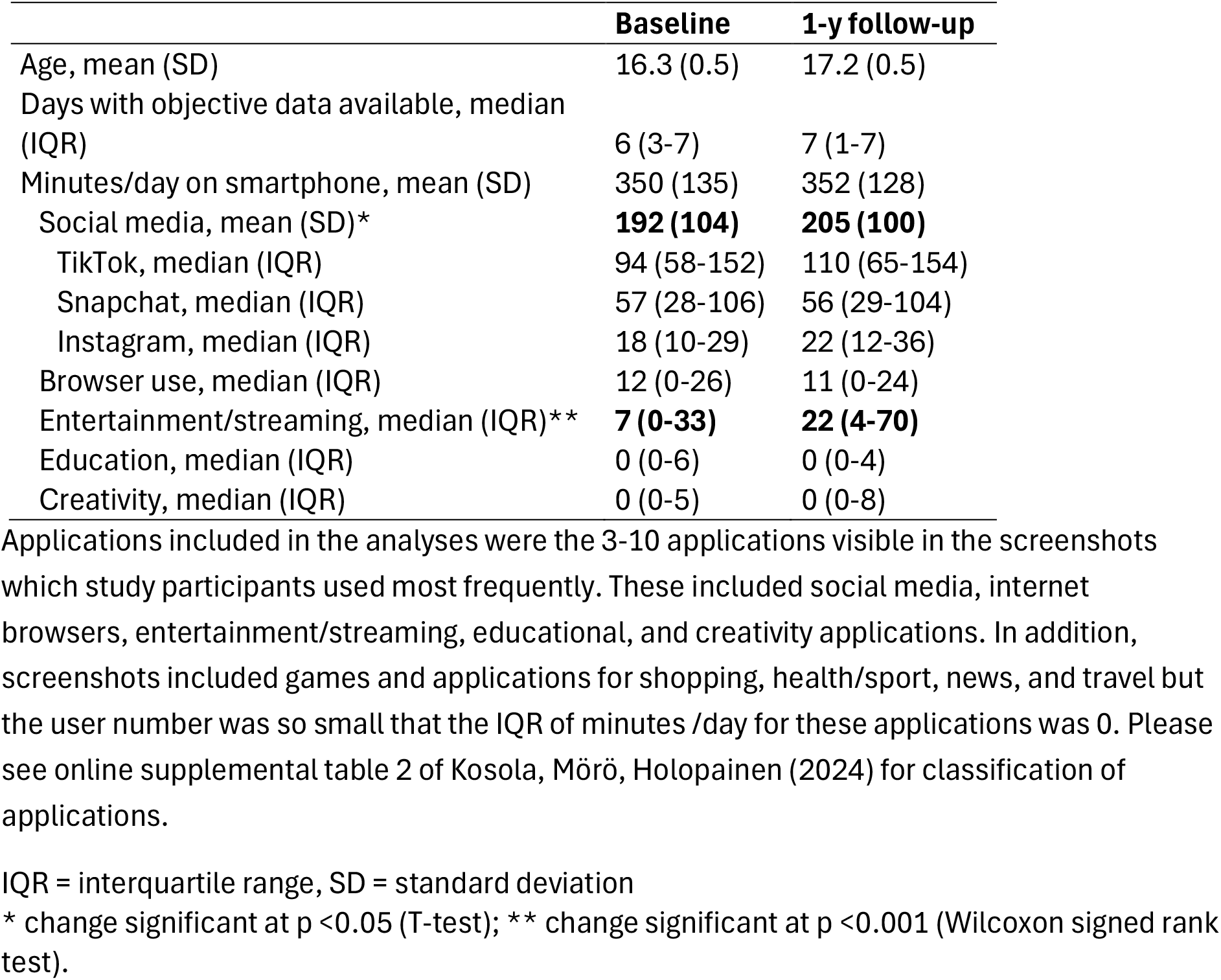
**Participant characteristics and objective smartphone use at baseline and one-year follow-up (n = 259 adolescent girls). Significant changes in bold**.

### Social media addiction

The Bergen Social Media Addiction Scale (BSMAS) was used to measure addictive behavior (18,19). BSMAS includes six statements with 5-point Likert scales (“very rarely” to “very often”). Possible scores range from 6 to 30, and higher points indicate higher risk of social media addiction. Scoring “often” or “very often” (i.e., 4 or 5 points) for at least four of the six items indicates addiction.

### Anxiety

The 7-item generalized anxiety disorder scale (GAD-7) was used to measure anxiety (20,21). Each item is scored on a 4-point Likert scale (“not at all” to “nearly every day”). Sum scores range from 0 to 21, and higher points indicate higher levels of anxiety. Scores 10-14 may indicate moderate anxiety and scores ≥15 severe anxiety.

### Body appreciation

The Body Appreciation Scale (BAS-2) was used to measure positive body image (22,23). The BAS-2 includes 10 items which are scored on a 5-point Likert scale (“never” to “always”). Total scores range from 10 to 50, and higher scores imply more positive body image.

### Tiredness, loneliness, and mood

Participants evaluated their current tiredness, loneliness, and mood on a visual analogue scale from 0 to 100 mm (24). Higher scores indicate better wellbeing.

### Statistical analysis

As descriptive statistics, frequencies are reported for days of screenshot data available and for prevalence of categorical social media addiction and possible anxiety. Means (with standard deviations, SD) and medians (with interquartile ranges, IQR) were used depending on data distribution. Adolescents with and without follow-up data and adolescents who sent screenshots and those who did not were compared in “sensitivity analyses”. Spearman correlation coefficients were calculated for continuous variables due to skewed distribution. We considered correlation coefficients of 0.10 to 0.29 weak, 0.30 to 0.49 medium, and 0.50 or higher strong (25). Mann-Whitney U test was used to compare groups based on potential social media addiction and use of different social media platforms. Wilcoxon signed rank tests were used for within-person comparisons of baseline and follow-up measures. To assess the associations of the most popular social media platforms with social media addiction and wellbeing outcomes, the highest user quartiles (25%) were compared with others to avoid small numbers in groups. IBM SPSS Statistics version 29.0 was used for analyses. P values below 0.05 were considered statistically significant.

### Ethics

This study involved human participants and was approved by the HUS Regional Committee on Medical Research Ethics (HUS/117/2022). Participants provided informed consent before participating in the study.

## Results

Of the 564 adolescents who completed the study survey and sent screenshots of their smartphone use at baseline, 259 (46 %) provided complete data including screenshots after one year of follow-up (Figure 1).

**Figure 1.**
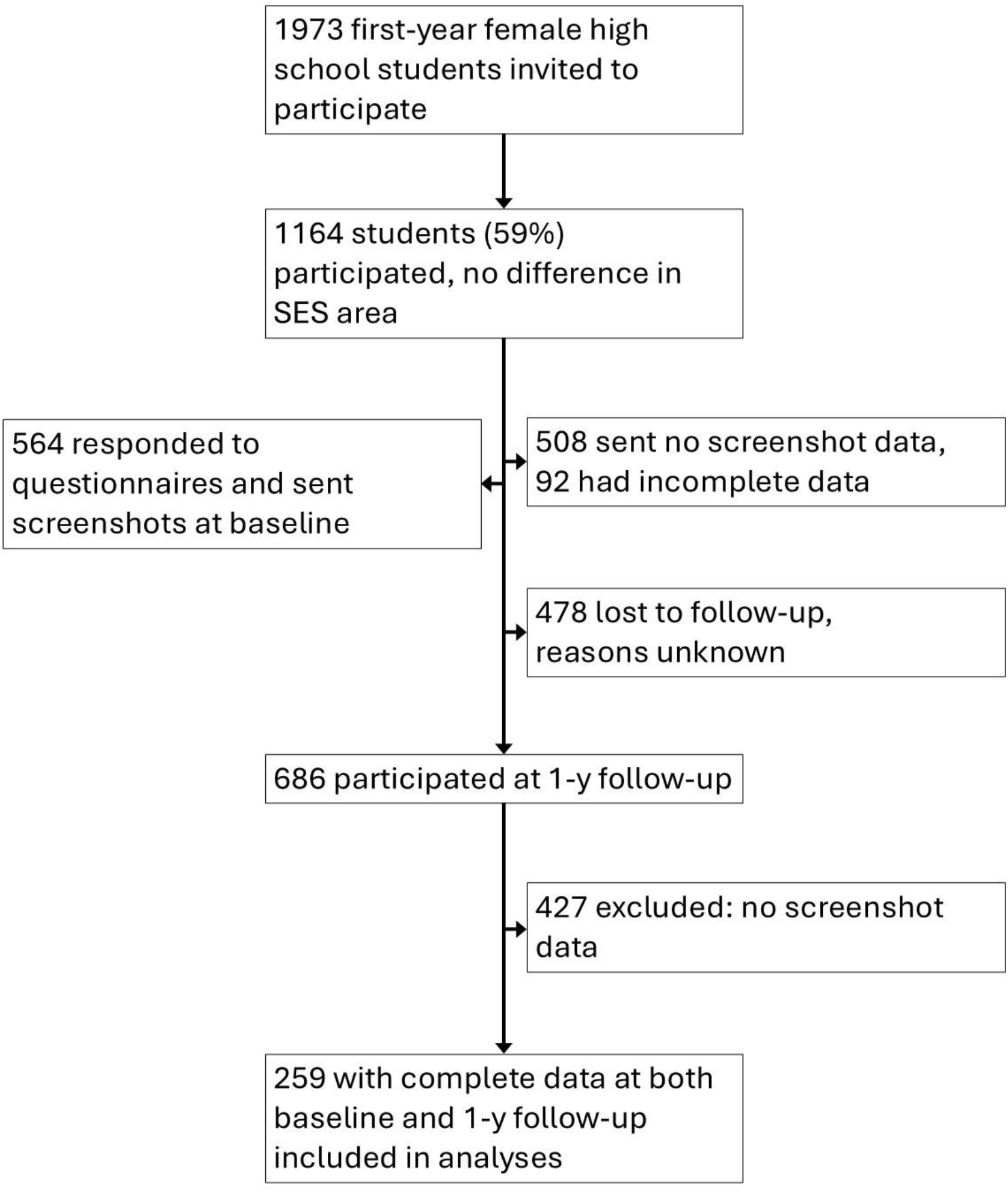
Flowchart depicting data collection.

The 259 participants included in longitudinal analyses did not differ in age, baseline prevalence of possible social media addiction, or baseline anxiety symptoms from their peers with no follow-up data. However, participants who sent screenshots at follow-up used social media less at baseline than did participants who did not send screenshots at follow-up: for the whole cohort at baseline, daily time on social media was 231 [121] minutes, among participants with objective data at follow-up, baseline social media use was 192 [104] minutes (p <0.001).

At baseline, participant age was 16.3 (0.5) years, and at one-year follow-up, 17.2 (0.5) years (Table 1). At follow-up, the time used on the 3-10 most popular applications in the screenshots explained 84% (IQR 76-92%) of time spent using smartphones. Social media use increased from 192 to 205 minutes per day (p =0.036) and streaming increased from 7 to 22 minutes per day (p <0.001). At baseline, 15% of the cohort had possible social media addiction, at one-year follow-up the prevalence was 17%.

In longitudinal analyses, social media addiction scores at baseline and follow-up showed a strong correlation (r =0.57, p <0.001). Overall, anxiety, body appreciation, and mood improved during the study period (Table 2), but baseline social media addiction scores were associated with increased anxiety (r=0.29, p<0.001), lower body appreciation (r=-0.15, p=0.022), and more loneliness (r=-0.20, p=0.001) at follow-up. When comparing groups with and without possible social media addiction at baseline, participants with addiction scored higher than their peers in anxiety, tiredness, and loneliness, and lower in mood (Table 3). Baseline anxiety was associated with social media addiction scores at follow-up (r=0.19, p=0.005). Baseline loneliness or body appreciation showed no association with social media addiction at follow-up.

**Table 2.**
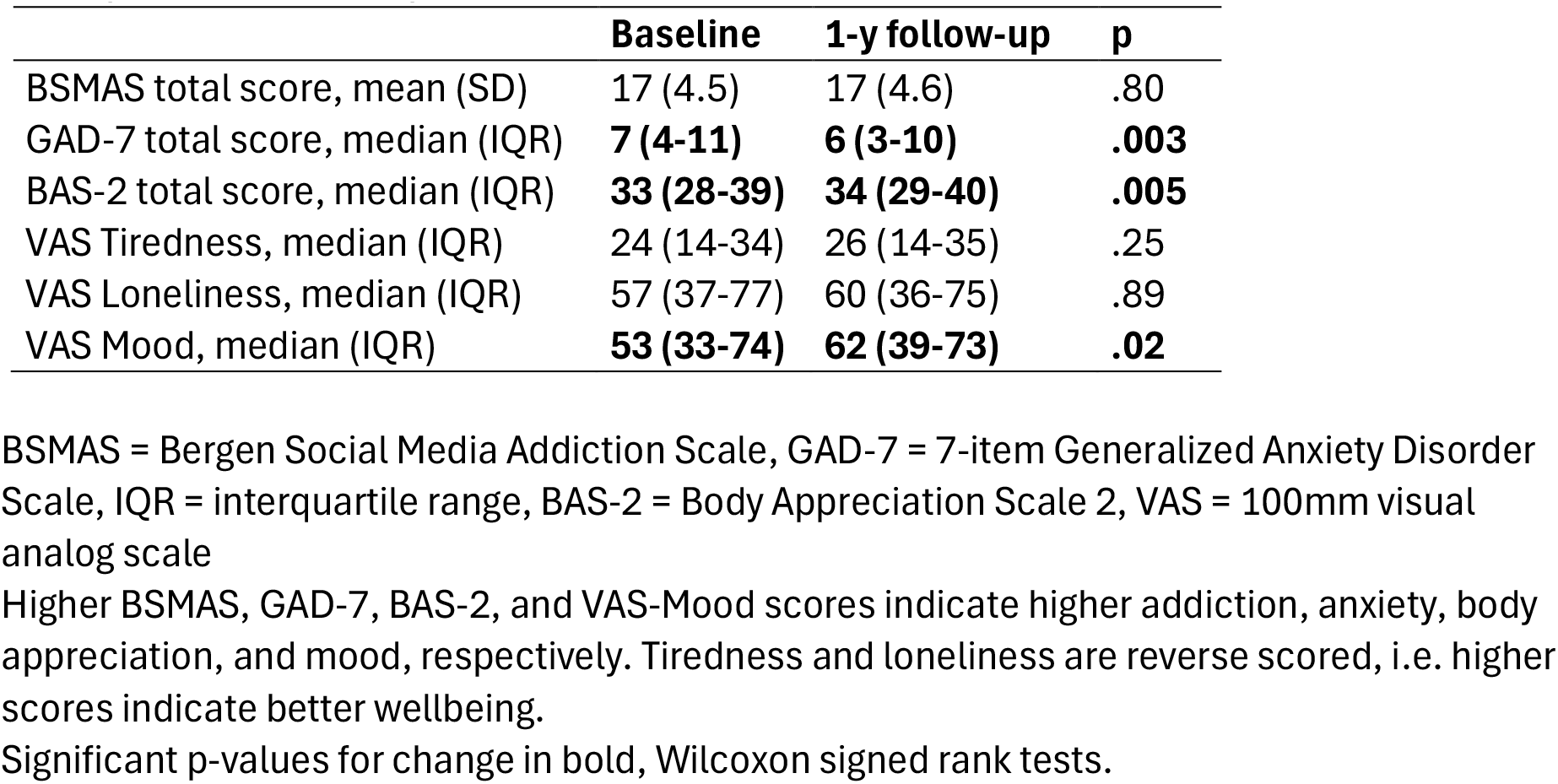
Social media addiction and wellbeing measures at baseline and one-year follow-up among 259 adolescent girls.

|  | **Baseline** | **1-y follow-up** | **p** |
| --- | --- | --- | --- |
| BSMAS total score, mean (SD) | 17 (4.5) | 17 (4.6) | .80 |
| GAD-7 total score, median (IQR) | **7 (4-11)** | **6 (3-10)** | **.003** |
| BAS-2 total score, median (IQR) | **33 (28-39)** | **34 (29-40)** | **.005** |
| VAS Tiredness, median (IQR) | 24 (14-34) | 26 (14-35) | .25 |
| VAS Loneliness, median (IQR) | 57 (37-77) | 60 (36-75) | .89 |
| VAS Mood, median (IQR) | **53 (33-74)** | **62 (39-73)** | **.02** |

BSMAS = Bergen Social Media Addiction Scale, GAD-7 = 7-item Generalized Anxiety Disorder Scale, IQR = interquartile range, BAS-2 = Body Appreciation Scale 2, VAS = 100mm visual analog scale
Higher BSMAS, GAD-7, BAS-2, and VAS-Mood scores indicate higher addiction, anxiety, body appreciation, and mood, respectively. Tiredness and loneliness are reverse scored, i.e. higher scores indicate better wellbeing.
Significant p-values for change in bold, Wilcoxon signed rank tests.

**Table 3.** Comparison of wellbeing measures after one year of follow-up between adolescent girls with and without potential social media addiction at baselines.

|  | **Addiction (n=38)** | **No addiction (n=221)** | **p** |
| --- | --- | --- | --- |
| GAD-7 total score, median (IQR) | **8 (6-15)** | **6 (3-10)** | **<0.001** |
| BAS-2 total score, median (IQR) | 33 (27-40) | 34 (29-40) | 0.47 |
| VAS Tiredness, median (IQR) | **14 (6-28)** | **27 (17-36)** | **<0.001** |
| VAS Loneliness, median (IQR) | **50 (29-66)** | **63 (38-77)** | **0.007** |
| VAS Mood, median (IQR) | **58 (31-69)** | **63 (40-73)** | **0.04** |

GAD-7 = 7-item Generalized Anxiety Disorder Scale, IQR = interquartile range, BAS-2 = Body Appreciation Scale 2, VAS = 100mm visual analog scale
Higher BSMAS, GAD-7, BAS-2, and VAS-Mood scores indicate higher addiction, anxiety, body appreciation, and mood, respectively. Tiredness and loneliness are reverse scored, i.e. higher scores indicate better wellbeing.
Significant p-values in bold, Mann-Whitney U test.

The three most popular social media platforms used were TikTok, Snapchat, and Instagram, respectively (Table 1). The highest quartile of TikTok users reported more social media addiction and lower body appreciation than did their peers (Table 4). The highest quartile of Snapchat users reported more social media addiction and tiredness than did their peers. No significant associations were found for Instagram use or anxiety.

**Table 4.** Comparison of wellbeing measures between the highest quartile of most popular social media platform users and others, data presented as medians (with interquartile ranges).

| Platform | BSMAS | GAD-7 | BAS-2 | Tiredness | Loneliness | Mood |
| --- | --- | --- | --- | --- | --- | --- |
| TikTok | **19 (16-21)** | 6 (4-11) | **32 (28-38)** | 21 (11-33) | 49 (32-74) | 59 (38-76) |
|  | **vs. 17 (14-20)\*\*** | vs. 6 (3-11) | **vs. 35 (29-40)\*\*** | vs. 26 (16-34) | vs. 65 (39-77) | vs. 63 (40-72) |
| Snapchat | **19 (15-21)** | 7 (4-10) | 34 (28-38) | **21 (13-32)** | 65 (38-78) | 60 (36-77) |
|  | **vs. 17 (14-20)\*\*** | vs. 6 (3-10) | vs. 34 (29-40) | **vs. 26 (15-35)\*** | vs. 61 (39-75) | vs. 63 (46-73) |

Significant differences between groups in bold. \*\*=significant at p <0.01 level, \*=significant at p <0.05 level.
Mann-Whitney U test.

## Discussion

In this longitudinal study, social media addiction was associated with increased anxiety, lower body appreciation, and increased loneliness one year later, while anxiety at baseline was associated with social media addiction at the end of follow-up. Objectively measured time spent using TikTok and Snapchat were associated with social media addiction. TikTok use was also associated with poorer body appreciation and Snapchat use was associated with increased tiredness.

We found a bidirectional association between anxiety and social media addiction, which may imply a vicious cycle. The associations we found between social media addiction and poor psychosocial wellbeing are in line with a recent review reporting results on younger adolescents from the longitudinal Adolescent Brain Cognitive Development Study (2). A systematic review of longitudinal studies on digital media use among 0-19-year-old children, however, found much smaller associations (26). Potential reasons for these diverging results may be explained by differences in study methodologies. Large age ranges may overlook stages of brain and social development which are relevant to the study context (1,27). Furthermore, due to the fast development of social media platforms, studies using data that was collected more than a decade ago may disregard the changes in operating mechanisms and use of psychological research in the development of social media algorithms (28,29). Finally, almost all studies still utilize self-reported data on social media use, although in our previous study, within subject time estimates and objectively collected data only showed a medium correlation (4).

According to the screenshots sent by our study participants, the most used social media platforms were TikTok, Snapchat, and Instagram, respectively. These platforms highlight the evolution of social media from brief text status posts into an image- and short video-based direction where filters can be used to enhance images even further and algorithms guide content. Unsurprisingly, the heaviest users of TikTok in our study reported lower body appreciation than did other study participants. TikTok has even been dubbed “SkinnyTok” for the amount of weight loss content it offers (30,31). The association of manipulated photos on social media with lower body image among adolescent girls is well-established (32,33). In a study by Sanzari et al, also undergraduate students in 2022 reported more body image problems and disordered eating habits than their peers did in 2015 (34). Most importantly, exposure to body positivity content was unable to protect from the harmful exposure to weight loss content.

In our study, social media addiction at baseline was associated with increased loneliness one year later, whereas loneliness at baseline showed no association with later social media addiction. In our cohort, both time spent on social media and time spent streaming content increased across follow-up, and passive time online has been previously associated with loneliness (12). Among the three most used social media platforms were TikTok, which emphasizes passive consumption of content, and Instagram, which is dedicated to cultivating personal brands and pressures to impress instead of promoting interaction between users (35). Other possible mechanisms for increased loneliness include fear of missing out and cyberbullying, which is prevalent on all popular social media platforms (36).

The heaviest users of Snapchat in our study reported more tiredness than their peers. Potential explanations include specific features such as streaks (consecutive days of exchanged snaps) and disappearing messages which create a sense of urgency and encourage 24/7 engagement. Snapchat users often manage their streaks late in the evening or even wake up during the night to check their feeds, which reduces sleep quantity and quality (37,38). Constant messaging may also lead to communication overload and emotional exhaustion (39).

To the best of our knowledge, this is the first study to utilize longitudinal, objective data on smartphone and social media use in combination with validated measures of social media addiction, anxiety, and body appreciation. The greatest limitation of our study is the attrition rate (54%) which means our study cohort may not be representative of the background population and the results should be interpreted with due caution. We carefully compared adolescents who participated at both baseline and follow-up with adolescents lost to follow-up and found that the retained participants used social media significantly less at baseline than did non-participants. It is plausible, however, that more intense social media use could have shown even stronger associations with psychosocial outcomes (40). It is thus unlikely that the study results would exaggerate the results. Unfortunately, we lack knowledge of the reasons for drop-out. We have no data on the age when study participants acquired their smartphone or when they accessed social media. The stability of the associations of social media addiction with psychosocial outcomes across follow-up increases the validity of previous cross-sectional results but in an observational study, no definitive conclusions on causal relationships can be drawn.

## Conclusion

The results of this longitudinal study suggest that adolescents who spend more time on popular social media platforms and develop social media addiction subsequently have poorer psychosocial wellbeing. Social media use and time spent watching streamed content increased from already high levels over time, possibly highlighting the addictive nature of several applications. Educating adolescents and caregivers on the potential harms of social media and increasing the availability of mental health services are insufficient measures as long as social media companies continue to develop more addictive platforms. Regulation is urgently needed to reduce the harm of social media.

## Data availability statement

Pseudonymous data are available upon reasonable request within the EU.

## Acknowledgements

The authors thank the members of the Youth Research Advisory Board, all study participants, and participating schools. Members of the Youth Research Advisory Board critically reviewed the study questionnaire. Medical student Ines Sederholm helped with recruitment of study participants and data acquisition.

This study was funded by the Foundation for Pediatric Research, the Finnish Medical Foundation, the Wihuri Foundation, the Department of Gynecology of the Helsinki University Hospital, The Society for Pediatric and Adolescent Gynecology, the Gyllenberg Foundation, the Olvi Foundation, and the Yrjö Jahnsson Foundation.

